# Re-emergence of dengue virus 3 genotype III in Cabo Verde, 2023

**DOI:** 10.1101/2024.02.29.24301580

**Authors:** Menilita dos Santos, Idrissa Dieng, Isaias Baptista Fernandes Varela, Kevin Sanders Da Rosa Carvalho, Domingos Dias Texeira, Ullardina Furtado, Liliane Hungria, Letícia Souza, Leidiza Tavares, Mamadou Aliou Barry, Cheikh Talla, Samba Niang Sagne, Ousmane Faye, Ndongo Dia, Cheikh Loucoubar, Oumar Faye, Amadou Alpha Sall, Boubacar Diallo, Moussa Moïse Diagne, Maria da Luz Lima, Abdourahmane Sow

## Abstract

We characterized 11 autochthonous dengue virus serotype 3 cases from Santiago and Fogo islands (Cabo Verde), 14 years after Cabo Verde’s latest noticed DENV outbreak involving this serotype. Identified viruses are closely related to Asian strains and falling into a clade distinct from known circulating West African DENV-3/Genotype III isolates.

## Background

Dengue virus (DENV) is a widely known and highly prevalent arthropod-borne virus globally (1). The virus is transmitted to human through the bite of infected Aedes mosquitoes, primarily *Aedes agypti* (1). The virus is divided into four antigenically and phylogenetically distinct serotypes (DENV 1-4). Each serotype is divided into genotypes, DENV-3 genotype III being one of the most widely distributed and transmissible strains (2). Cabo Verde, a volcanic archipelago nation off the coast of West Africa, spans 40323 km2 and is located approximately 550km off Senegal’s coast.

The archipelago consists of 10 islands, nine of which are inhabited with approximately 491.233 inhabitants (3) (Figure 1). Despite no report of recent arboviruses circulation, arbovirus vectors’ are abundant (4) and the country has experienced large DENV (4) and ZIKV (5) epidemics in the past.

**Figure 1:**
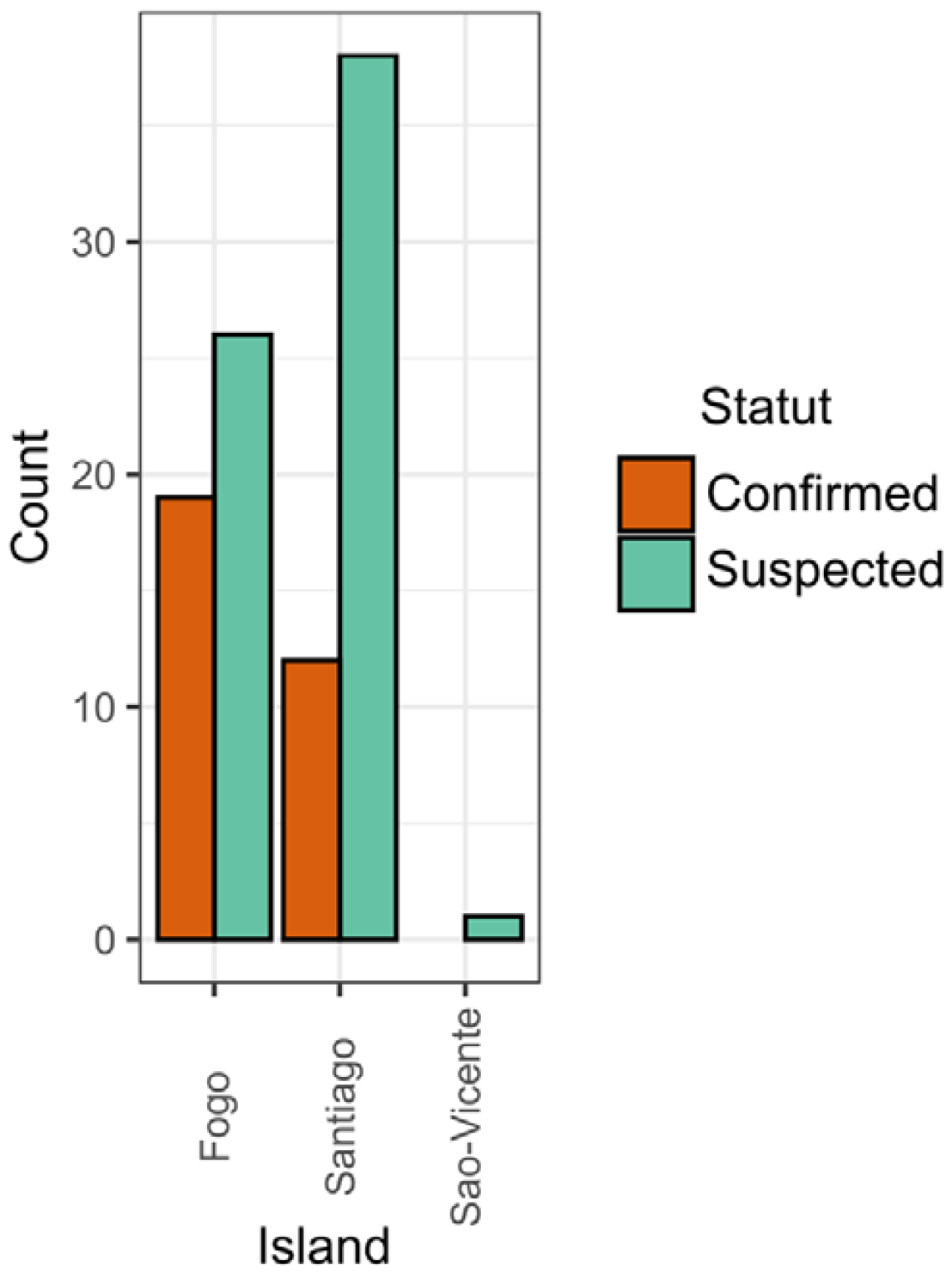
Barplot showing the number of confirmed and suspected DENV cases by island and covering the study period

### The study

On April 20 2022, the Africa CDC launched RILSNET program(), a project utilizing the Syndromic Sentinel Surveillance Network in Senegal (4S Network) which has helped detect over 30 outbreaks since 2015 (data no shown), as a model to enhance febrile illnesses surveillance in West Africa. The first cohort included Cabo Verbe, Gambia, and Mauritania.

The project support selecting countries by providing reagents, consumables, laboratory and epidemiological training to set up sentinel surveillance of febrile illnesses etiologies including DENV. In this context a team of Institut Pasteur of Dakar (IPD) was deployed from October 22, 2023 to November 05 2023 to Santiago and Sal islands respectively to train laboratory technicians, epidemiologists and health care givers around syndromic surveillance of arboviruses and respiratory viruses.

On November 2023 through the newly implemented sentinel sites in Cabo verde, two autochtonous DENV cases were detected in Santiago. Serum samples were sent to the Institut Pasteur de Dakar (IPD)’s regional reference laboratory for confirmatory tests, where they were extracted using the Qiagen viral RNA mini kit before a panDENV RT-qPCR as previously described (6). Tests confirmed a DENV epidemic in Cabo Verde on November 08 2023, prompting the Cabo Verde Ministry of Health to establish a specific surveillance system. By November 2023, xx confirmed DENV cases were found in the hospital’s virology lab of the National Public Health Institute (NPHI), following the same protocol used at IPD.

On November 28, 2023, NPHI and IPD partnered to conduct real-time viral serotyping and genomic surveillance at virology lab in Cabo Verde. Nucleic acid extraction was performed on confirmed RT-qPCR DENV positive samples and RNA extracts were retrospectively subjected to molecular serotyping using Tib MolBiol Dengue typing kit (**Cat**-No. 40-0700-24) as recommended by the manufacturer. All thirty-one tested samples were positive for DENV-3. Eleven samples with the best Ct values (*Table 1*) were selected for cDNA synthesis using reverse transcriptase LunaScript (NebNext). An amplicon based tilling PCR strategy followed by whole genome sequencing on a Oxford Nanopore Technology MinION MK1C platform was then performed as previously described (6). Nearly complete genomes were obtained for all the eleven sequenced DENV samples, all exhibiting good coverage values above 89 % (*Table 1; Figure S2*),with background dataset containing described DENV-3 genotypes and closely related worldwide sequences. Based on a dataset containing described DENV-3 genotypes and closely related sequences, Maximum Likelihood (ML) trees were built for the purpose of viral genotyping and decipher relationship with global strains.

**Table 1:** Epidemiological data and sequencing statistics of the 11 DENV-3 genome sequences obtained as part of this study. Used sample IDs was anonymized and cant be linked to any physical individual.

| Sample ID | Island of provenance | Date of Collection | Ct value | Number of reads mapped to reference | Depth | Coverage |
| --- | --- | --- | --- | --- | --- | --- |
| 003 | Santiago | November 2023 | 25.21 | 32418 | 2535.8 | 93.3% |
| 005 | Santiago | November 2023 | 28.26 | 10186 | 837.9 | 89.0% |
| 011 | Santiago | November 2023 | 25.07 | 20048 | 1409.3 | 96.4% |
| 032 | Fogo | November 2023 | 20.31 | 34715 | 2539.6 | 96.4% |
| 034 | Fogo | November 2023 | 22.18 | 29226 | 1966.0 | 96.5% |
| 037 | Fogo | November 2023 | 26.34 | 31343 | 2254.8 | 96.8% |
| 041 | Fogo | November 2023 | 24.21 | 32951 | 2297.4 | 96.8% |
| 046 | Fogo | November 2023 | 20.45 | 39114 | 2651.5 | 96.5% |
| 048 | Fogo | November 2023 | 22.25 | 28409 | 1955.0 | 96.8% |
| 057 | Santiago | November 2023 | 22.22 | 26884 | 1860.6 | 96.5% |
| 060 | Santiago | November 2023 | 19.06 | 33224 | 2213.7 | 96.5% |

Our results showed that since the notification of the first case up to 30 November 65 suspected DENV cases samples were received and subjected to viral RNA detection at NPHI virology lab; 31 out of them were DENV RNA+ with Fogo island exhibiting the higher number positive (n = 19) followed by Santiago (n = 12) (Figure 1 ; Figure 2). None DENV+ were obtained in Sao-Vincente.

**Figure 2:**
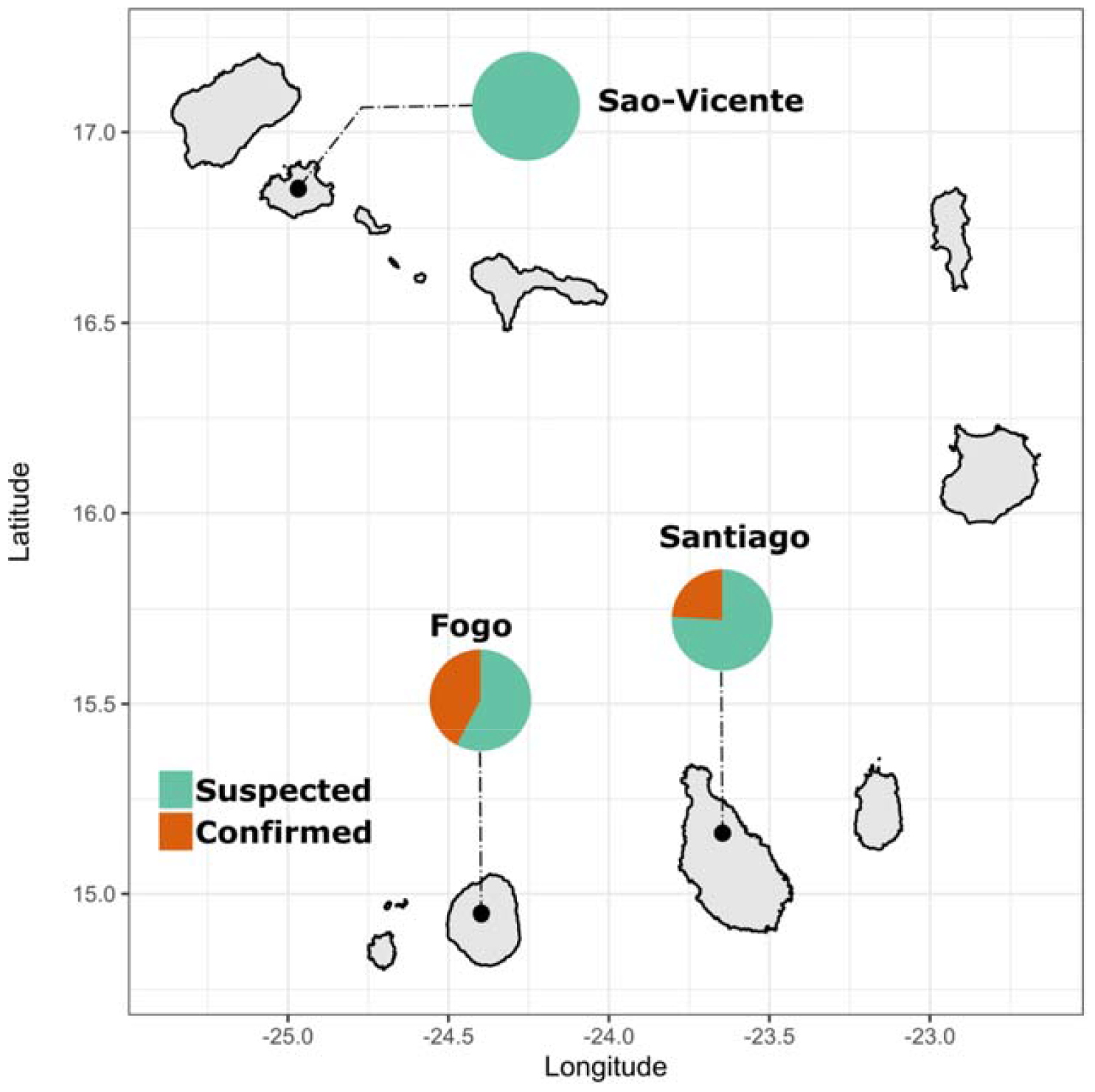
Map showing the spatial distribution of suspected and confirmed DENV-3 samples in Cabo Verde in November 2023. The pie charts size is not proportional to the number of cases; number of cases are represented on figure xx. Orange color represent the number of confirmed cases by RT-qPCR while green color the number of suspected cases shipped to the Institut National de Saude Publica (INSP) virology lab.

ML genotyping tree (*Figure 3*) confirmed that sequences strains belong to Genotype III, which was responsible for the Cabo Verde 2009 epidemic (7)(*ref*). Additionally, the global tree (*Figure S1*) confirmed that circulating DENV-3/GIII strains in Cabo Verde branched with those in Asia, forming a distinct clade with previously known strains in West Africa (6,8).

**Figure 3:**
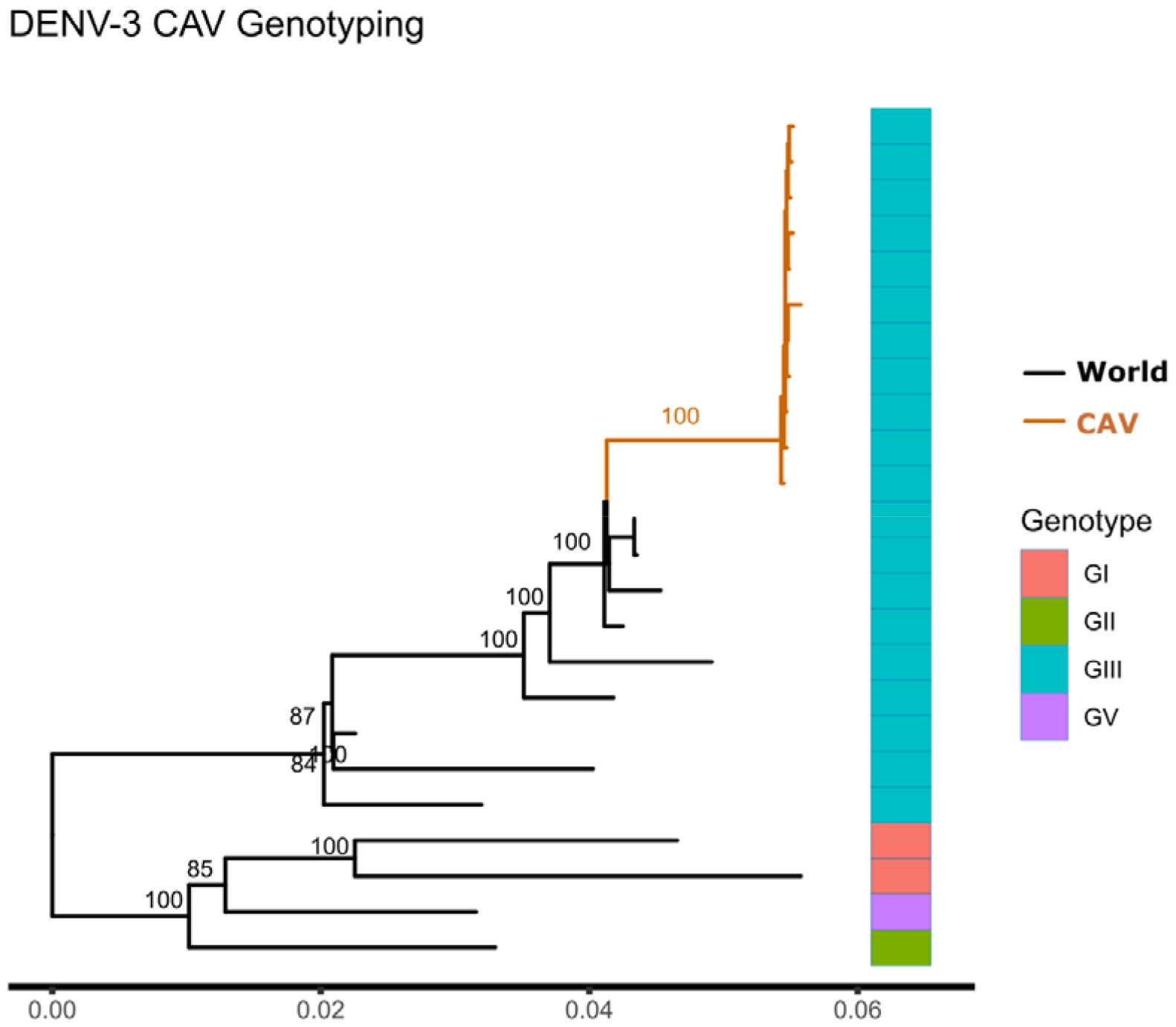
Maximum likelihood ML tree showing the genotype of obtained Cabo Verdien DENV-3 sequences. Tree was made using nearly complete genome sequences; 1000 replicates were performed for robustness. Cluster from Cabo Verde is colored in Orange. Heatmap depicted the genotype of used genome sequences.

The wider dissemination of the newly detected DENV-3/GIII strains in Cabo Verde may be concerning since tourism is the primary drive of the archipelagos economy (9), and naïve travelers could drive unprecedented viral diffusion. In another hand previous entomological studies demonstrated the high vector competency of local mosquitoes for DENV-3 (10) while Breteau Index above epidemic risk was reported in 15 out of 22 expected municipalities around the archipelagos (4).

We herein emphasized the importance of the newly implemented arbovirus surveillance through syndromic approaches in Cabo Verde. Additionally, a large-scale deployment of this sentinel surveillance model around the country is key to improve local infectious disease surveillance program.

## Data Availability

All data produced in the present study are available upon reasonable request to the authors

## Ethical considerations

The Cabo Verdien National Ethical Committee of the Ministry of Health approved the surveillance protocol which lead to the obtention of human sera as less than minimal risk research, and written consent were not required. Throughout the study, the database was shared with le Service Integrated Surveillance and Riposte of Cabo Verde and the Ministry of Health for appropriate public health action.

## Funding

This work was funded by Bill and Melinda Gates Foundation under grant (INV-050589) and the Africa Pathogen Genomics Initiative (CARES grant 4306-22-EIPHLSS-GENOMICS).

## Acknowledgments

We would like to thanks Marc Bulterys from Bill and Melinda Gates Foundation and to convey warm thanks to all the health workers of Cabo Verde archipelagos, workers of the INSP virology laboratory at Augustino Neto Hospital.

---

**Figure S1:**
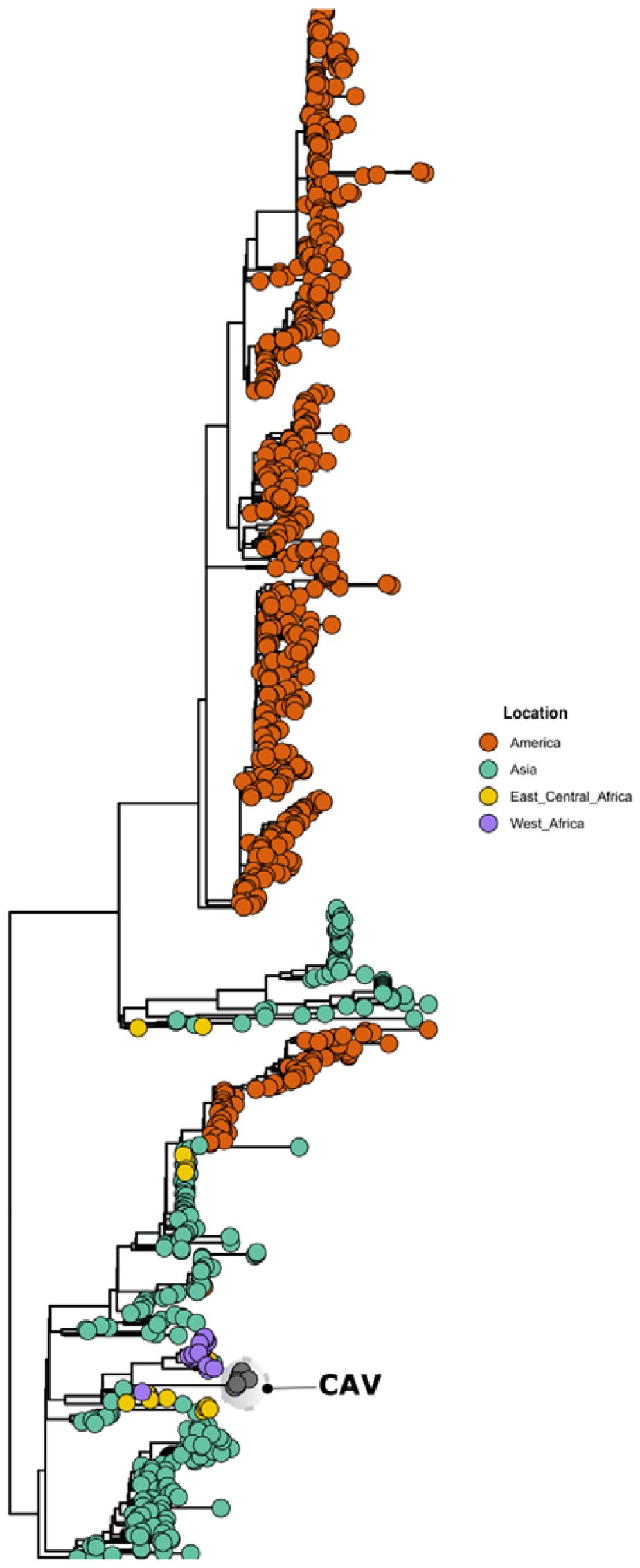
Maximum Likelihood tree accounting 998 Global DENV-3/GIII sequences supplemented with 11 newly generated nearly complete Cabo Verdien sequences highlighted by the grey circle.

**Figure S2:**
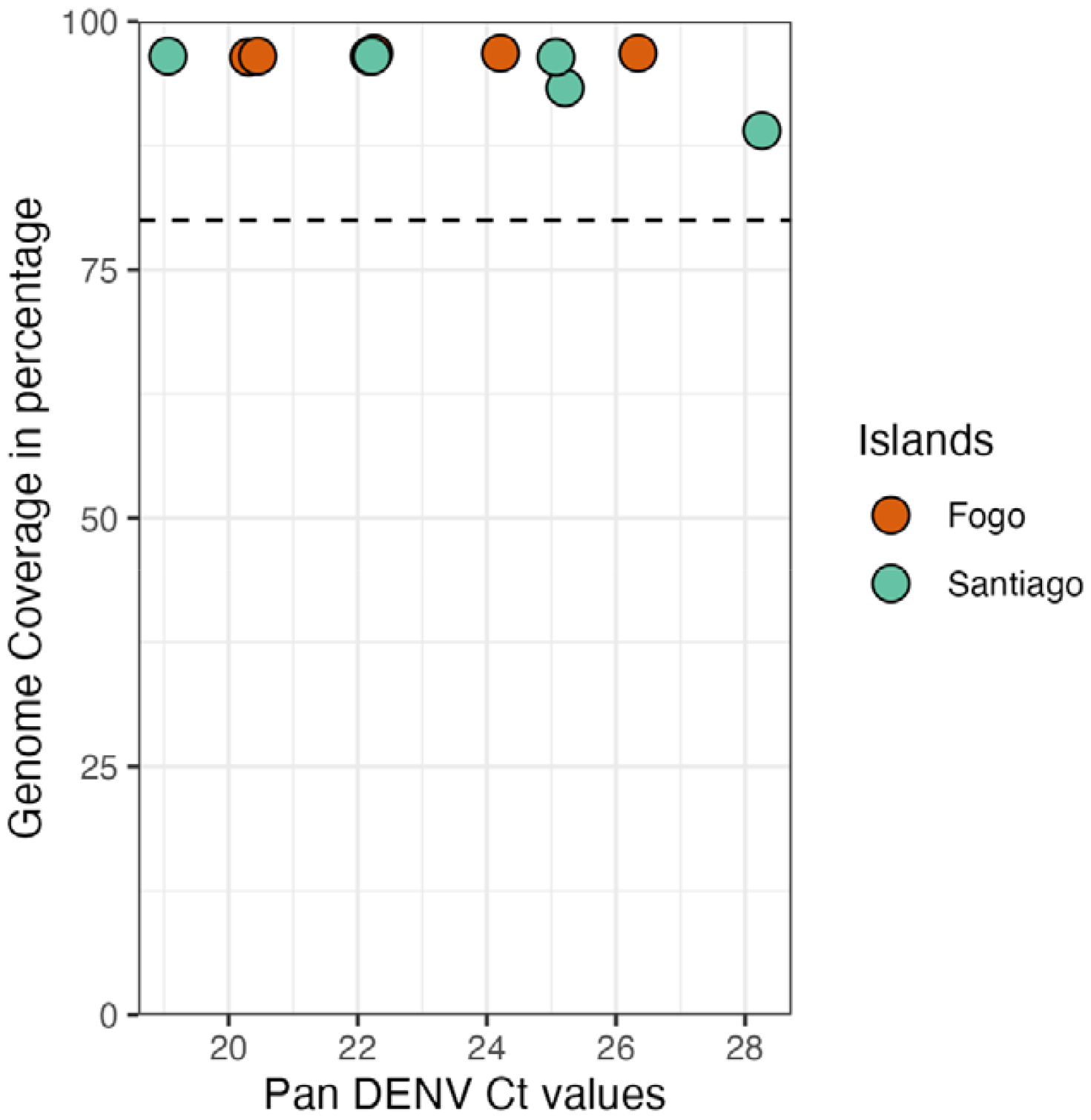
Relationship between panDENV Ct values and genome coverage of sequenced DENV RT-qPCR+ samples during this study.

## Notes

### Competing Interest Statement

The authors have declared no competing interest.

### Author Declarations

The Cabo Verdien National Ethical Committee of the Ministry of Health approved the surveillance protocol which lead to the obtention of human sera as less than minimal risk research, and written consent were not required.

